# Integrating Longitudinal Cohorts for Population Health Research: Mainland and Island-based Puerto Rican Diabetes Study

**DOI:** 10.64898/2026.10.08.26365065

**Authors:** Andreia Martinho, Martha Tamez, Josiemer Mattei, Elizabeth Petit, Sabrina E. Noel, Bayan Tuffaha, Katherine L. Tucker, Jose M. Ordovas, Abani Patra, Rebecca Batorsky

**Affiliations:** Tufts Institute for Artificial Intelligence, Tufts University, Joyce Cummings Center, 177 College Avenue, Medford, MA 02155; Harvard T.H. Chan School of Public Health, 655 Huntington Ave, Boston, MA 02115; Department of Biomedical and Nutritional Sciences, University of Massachusetts, Lowell, MA, USA; Jean Mayer USDA Human Nutrition Research Center on Aging at Tufts University, 711 Washington Street, Boston, MA 02111

## Abstract

**Background:** Population research increasingly relies on integrating data across studies to understand patterns of health and disease at broader scales. Such integration expands sample size and supports multimodal analyses, but independent cohort studies are often not directly comparable due to differences in instruments, timing, and measurement structure. Data harmonization addresses this challenge by transforming heterogeneous variables into comparable measures, allowing datasets collected under different protocols to be analyzed jointly, but published harmonization efforts seldom include an analysis conducted on the integrated dataset itself.

**Methods:** We developed an integrated dataset, combining two longitudinal cohorts, the Boston Puerto Rican Health Study (BPRHS) and the Puerto Rico Observational Study of Psychosocial, Environmental, and Chronic Disease Trends (PROSPECT). Comparability between variables was established through an explicit framework distinguishing availability from construct comparability, and the evidence supporting comparability is reported by class of feature, including tests of measurement invariance and differential test functioning for the multi-item psychosocial scales. To support transparency and reuse, we provide an open-source Python pipeline and structured metadata describing variable alignment and transformations. As a worked example of downstream use, we conducted a Random Forest analysis predicting diabetes status.

**Results:** The integrated dataset comprises 539 harmonized variables across four BPRHS visits and two PROSPECT visits, each documented with its source variable, question wording, and transformation. In the worked example, the integrated dataset retained predictive signal relative to cohort-specific models and improved performance for the smaller cohort.

**Conclusions:** These findings illustrate how transparent, documented cross-cohort integration can support predictive population health research while making the basis for comparability explicit.

## Introduction

Population research increasingly integrates data across studies to understand patterns of health and disease on broader scales (1–3). Such integration allows researchers to increase statistical power, study related populations across contexts, and support more generalizable findings (4, 5). However, independent cohort studies are rarely directly comparable. Differences in instruments, measurement scales, timing of collection, and study design often prevent comparable evidence from being generated across studies. As a result, the potential benefits of cross-cohort integration remain difficult to realize in practice.

Data harmonization addresses this challenge by transforming heterogeneous variables into comparable measures, allowing datasets collected under different protocols to be analyzed jointly (3, 6–9).

Much of this work has focused on pooled analyses across cohorts, often integrating studies from different regions to enhance demographic diversity and enable cross-population inference (4, 10–14). Several studies also describe practical challenges encountered during harmonization, including differences in sampling, study design, and measurement instruments, and report quality-control checks such as comparisons of variable distributions across timepoints (7, 10–13). Beyond these applied studies, methodological guidance has been proposed to support retrospective harmonization and survey integration, including recommended validation procedures and consistency checks (3, 6, 9, 15).

These contributions demonstrate how datasets can be made comparable, but seldom include an analysis conducted on the integrated dataset itself. Documented implementations and worked examples remain uncommon. Yet such examples are valuable to harmonization research, since applying an integrated dataset in a concrete test case analysis illustrates what the data can support.

In this study, we developed and documented an integrated dataset combining two longitudinal population cohorts: the Boston Puerto Rican Health Study (BPRHS) (16) and the Puerto Rico Observational Study of Psychosocial, Environmental, and Chronic Disease Trends (PROSPECT) (17). These cohorts provide a compelling setting due to their complementary contexts, demographic diversity, and relevance to chronic disease research, and illustrate challenges commonly encountered when integrating minority-focused longitudinal health datasets.

Whereas our primary contribution is the harmonized BPRHS/PROSPECT resource, an open-source integration pipeline and structured metadata documenting variable definitions, transformations, data quality, and availability across cohorts and timepoints, we also provide a worked example of a downstream analysis, predicting diabetes status -a prevalent condition in both cohorts, shaped by lifestyle, biological, and socioeconomic factors - with a Random Forest model. By following the harmonized data, from variable selection through model specification and to evaluation, this study contributes a documented approach that may inform future cross-cohort integration of similar population studies.

## Methods

### Overview of the Source Cohorts

The BPRHS (16) and PROSPECT (17) are longitudinal cohort studies of Puerto Rican adults conducted in distinct geographic contexts. BPRHS follows approximately 1500 participants residing in the greater Boston area and collects socio-economic, nutritional, clinical, and metabolomic information. PROSPECT follows approximately 1850 participants residing in Puerto Rico and collects socio-economic, psychosocial, nutritional, lifestyle, and health data. Participants in BPRHS are older on average (mean age approximately 57–69 years across visits), with lower educational attainment (modal category: 9th–12th grade or GED) and lower household income, and exhibit a higher glycemic burden. In contrast, PROSPECT participants are younger (mean age approximately 53–58 years across visits), have higher educational attainment (modal category: BA or BS degree), similar levels of adiposity (BMI approximately 31–32 kg/m²), and more favorable glycemic indices (Table 1). Differences between the cohorts and visits that bear directly on harmonization, including the calendar years of data collection, the intervals between visits, the languages of administration, the instruments administered and their recall periods, and examples of differences in question wording, response categories and clinical measurement protocols, are summarized in SI

**Table 1.** Characteristics of the BPRHS and PROSPECT cohorts at each study visit. Sample size is the number of participants with a recorded visit date at that study visit; the number completing the food frequency questionnaire (FFQ) is reported separately, as the FFQ was administered as a separate contact, and is the number of participants with an FFQ processed into nutrient intakes for that visit (at BPRHS visit 3, the FFQ count exceeds clinic attendance because the 5-year FFQ file includes 396 records whose identifiers could not be linked to a visit 3 clinic participant). Characteristics comprise age, sex, household income (reported as median (IQR) of annual household income in BPRHS, and as the modal income bracket, n (%), in PROSPECT, where income was collected in categories.), education, body mass index (BMI), glucose, and glycated hemoglobin. Continuous variables are summarized as mean (SD) and categorical variables as n (%), computed among participants attending the visit who have a valid value for that variable.

| **Cohort** | **BPRHS** |  |  |  | **PROSPECT** |  |
| --- | --- | --- | --- | --- | --- | --- |
| **Visits** | **V1** | **V2** | **V3** | **V4** | **V1** | **V2** |
| Sample (attended visit), n | 1502 | 1258 | 961 | 573 | 1737 | 525 |
| Sample size (completed ffq), n | 1491 | 1244 | 1256 | 578 | 1149 | 103 |
| Age, mean (SD) | 57 (7.60) | 59 (7.67) | 63 (7.65) | 69 (7.48) | 53 (11.71) | 58 (10.93) |
| Female, n (%) | 1059 (70.5%) | 905 (71.9%) | 693 (72.1%) | 426 (74.3%) | 1286 (74.1%) | 369 (70.3%) |
| Age, Female, mean (SD) | 57 (7.45) | 59 (7.48) | 63 (7.49) | 69 (7.37) | 53 (11.72) | 58 (11.09) |
| Age, Male, mean (SD) | 57 (7.93) | 60 (8.12) | 63 (8.06) | 68 (7.72) | 54 (11.69) | 58 (10.58) |
| Household Median USD (IQR) (%) | 11063 (10758) | 11124 (7751) | 11520 (5130) | 15600 (13059) | 0-10K 459 (27.7%) | 0-10K 129 (26.4%) |
| Education, mode, n (%) | 9th–12th or GED, 571 (38.2%) | 9th–12th or GED, 477 (38.0%) | 9th–12th or GED, 362 (38.0%) | – | BA or BS degree, 489 (28.3%) | BA or BS degree, 148 (28.4%) |
| BMI, mean (SD) | 31.9 (6.66) | 31.8 (6.62) | 31.1 (6.37) | 31.1 (6.12) | 31.4 (7.41) | 31.6 (7.51) |
| Glucose (mg/dl), mean (SD) | 120.26 (52.60) | 116.86 (49.05) | 117.50 (53.08) | 130.61 (51.05) | 106.23 (42.51) | 106.76 (42.36) |
| Glycated hemoglobin (%), mean (SD) | 6.96 (1.76) | 6.83 (1.55) | 6.69 (1.47) | – | 6.16 (1.41) | 6.11 (1.30) |

Table 3. For the features harmonized across cohorts, the recall periods coincided (for example, past week for the CES-D, past month for the Perceived Stress Scale, and both the past 12 months and the lifetime for the Perceived Discrimination Questionnaire, which asks each item for both periods).

### Retrospective Harmonization of the BPRHS and PROSPECT datasets

In order to integrate the BPRHS and PROSPECT datasets, we relied on data harmonization. This process can take place prospectively during study design, or retrospectively after data collection (6). We used retrospective harmonization, since the data had already been collected, and a flexible approach, in which variables representing comparable constructs were transformed to a common operational definition despite differences in wording, scale, or collection structure, with the aim of achieving inferential equivalence (6). The harmonization endeavor followed a three-step approach, including (i) *Preparatory*; (ii) *Implementation*; and (iii) *Post-harmonization and validation* (6). In the (i) *Preparatory* phase, we established a collaboration with the institutions involved with both BPRHS and PROSPECT research, namely University of Massachusetts Lowell and Harvard University. The raw data sources consist of a main questionnaire covering a wide range of topics, from health to socio-economics, and a food frequency questionnaire (FFQ) (18), processed to nutrients with the Nutrition Data System for Research (NDSR) [version 5.0_35]. This collaboration extended beyond data access and entailed ongoing support and guidance on the datasets: questions that the source documentation left unresolved, such as the meaning of a response option or how an item was administered, were referred to investigators and data managers of each cohort, and features for which no common definition could be agreed were not harmonized. For reasons of governance, compliance, and transparency, and to mitigate the potential risks associated with revising the Data Usage Agreements (DUA) across institutions, we decided not to host the harmonized data, instead, we make available the information about the input datasets (from both BPRHS and PROSPECT), the harmonization code, and the metadata (SI table 1) as well as a synthetic dataset that can be used to test the harmonization pipeline. This approach fosters transparency, as researchers can directly rerun the harmonization pipeline on the original inputs, thereby reproducing the integrated harmonized dataset.

In the (ii) *Implementation* phase, we defined the features that would be harmonized. We first identified groups of features within the different modalities of data (Health, Social Determinants of Health (SDOH), and Nutrition (FFQ)) across the BPRHS and PROSPECT datasets, through the analysis of the codebooks. The Health and SDOH variables were derived from the main questionnaire, whereas the FFQ variables were derived from the FFQ questionnaire. Most of the variables were straightforward to classify into different groups (e.g., medical diagnoses, medications, supplements, etc.), as we relied on the previous classification provided by the codebooks of BPRHS and PROSPECT. For the SDOH, we also relied on the NIH PhenX Social Determinants of Health collection, which consists of data protocols to help measure individual as well as structural factors that shape behaviors and health outcomes to further identify the structural and individual groups of features within the data (e.g., cleanliness and healthiness of the neighborhood, household income).

Within each group of features, we subsequently screened for identical features across the different time points within both BPRHS and PROSPECT. Our framework to establish comparability between variables comprised two dimensions: (i) Availability and (ii) Construct Comparability. The first dimension (Availability) entailed checking the codebooks to establish whether a construct was present in both cohorts, rather than simply matching variable names, since equivalent constructs were frequently recorded under different names. Features present in both cohorts, even if at a limited number of timepoints, were retained, with their availability across BPRHS V1-V4 and PROSPECT V1-V2 recorded in the metadata (SI table 1). This dimension resulted in two outcomes with respect to variables: (A) available in both cohorts and (B) not available in both cohorts.

The second dimension (Construct Comparability) was applied to subset (A), features available in both cohorts. By construct comparability we mean that the source variables measure the same construct and can be brought to a common operational definition. This is a necessary condition for inferential equivalence, the aim of harmonization, but not the same thing: inferential equivalence is the stronger property that analyses of the harmonized variable support the same inferences in both cohorts, and we assessed it empirically where the data permitted (see Quality Control). We compared the variable definitions documented in the codebooks to assess whether a transformation was needed to bring the sources to a common operational definition. Where the definitions already coincided, the feature was harmonized without transformation (for example, hmz_health_ant_bmi, which relates to body mass index); where the sources captured the same construct but differed in their operationalization, we defined the transformations required to bring them to a common operational definition (for example, hmz_health_ant_avg_waist, which relates to average waist circumference, for which BPRHS values were converted from centimeters to inches); and where the sources did not capture the same construct, or where no transformation could bring them to a common operational definition without altering what the variable captured, the feature was not harmonized. One feature, the number of alcoholic drinks consumed per drinking day, was excluded following this criterion: both questionnaires asked the same question with the same standard-drink definitions (a 12 oz beer, a 4 oz glass of wine, or 1 oz of liquor), but PROSPECT responses clustered at multiples of 12 (12, 24, 48 and 72), indicating that a subset of respondents had reported ounces rather than number of drinks, which cannot be distinguished from genuine counts in the data. This dimension therefore resulted in three different outcomes when it comes to the variables: (C) harmonizable without transformation, (D) harmonizable with transformation, and (E) not harmonizable.

Alongside the harmonization of individual features, the two cohorts required a common temporal reference. Visit numbers are not comparable across cohorts, since the cohorts were followed over different calendar periods and at different nominal intervals, and the observed intervals between visits varied between participants. We therefore derived a common temporal measure, hmz_days_since_visit_1, defined for each participant as the number of days between the date of a given visit and the date of that participant’s first visit, so that visit 1 takes the value zero in both cohorts. This variable places both cohorts on a single time axis expressed in days since study entry, and allows analyses to be conducted on elapsed time rather than on visit number.

The transformations varied in complexity, understood here as the extent to which the transformation followed directly from the variable definitions rather than requiring a judgment that the documentation alone could not settle. Straightforward transformations (syntactic harmonization) were fully determined by the variable definitions, and concerned features that differed only in the units or names in which they were recorded. These were resolved by renaming features, converting units of measurement, or computing ratios. For example, average body weight (hmz_health_ant_wt_avg) was measured in the same way in both cohorts and required only converting BPRHS values from kilograms to pounds. Moderately complex transformations (semantic harmonization) were not fully determined by the response options, and required a decision about how the two sets of responses should be aligned. For example, perceived neighborhood violence (hmz_sdoh_nfa_violence) was recorded in both cohorts on a five-point agreement scale, but the two questions were phrased in opposite terms: BPRHS asked participants whether they agreed that violence is a problem in their neighborhood, while PROSPECT asked whether they agreed that it is not. Matching the response values directly would therefore have reversed the direction of the measure. We adopted the PROSPECT phrasing as the harmonized definition and reverse-coded the BPRHS responses, so that higher values indicate greater perceived violence. The most difficult transformations concerned features that neither cohort recorded directly, and which had to be constructed from the underlying responses according to a definition we established. For example, neither questionnaire asked whether a participant held public health insurance (hmz_sdoh_public_insurance). Instead, participants were asked to indicate which insurance programs covered them, from lists that differed between the cohorts. Deriving the harmonized variable therefore required defining, for each cohort’s list, which of the named programs constitute public insurance, and treating a participant as covered if any of those was selected (SI table 1).

These transformations were coded in Python and integrated into a harmonization pipeline that is available on https://github.com/rbatorsky/diabetes-cohort-harmonization, for the transparency and reusability of this research, and are also well-documented in the metadata (SI table 1) .The integrated dataset includes a mix of individual and structural SDOH features, related to household composition, neighborhood food and activity, social and community support, access to health, perceived discrimination, food security, income, and migration history (age and sex were considered both SDOH and Health features); health features related to laboratory assays, anthropometrics, medical diagnoses, medical family history, health behaviors, perceived stress scale, and depression scale; and also FFQ features related to diet type, meal place, supplements, and nutrients (Table 3). All harmonized features were renamed, and a prefix hmz_ was added, standing for harmonization. Additional prefixes were added to identify the different modalities of data, namely sdoh_ for social determinants of health, health_ for the health features, and ffq_ for nutrition (food frequency questionnaire) features, as well as for the different groups of variables within each modality. For example, hc_ for household composition, med_ for medical diagnoses, and sup_dur for supplements use duration.

To support the use of the integrated dataset and provide transparency regarding the transformations in the pipeline, the metadata file (SI Table 1) contains detailed information about each harmonized feature, including the feature name, data modality (Health, SDOH, or FFQ)^1^, feature type (numerical or categorical), definition, and any transformations applied. It also documents the presence of each feature across different BPRHS timepoints (V1, V2, V3, and V4) and PROSPECT timepoints (V1 and V2). Additionally, the metadata identifies the original source features from which each harmonized feature was derived, including their definitions and the source files for both BPRHS and PROSPECT.

In the (iii) Post-harmonization and validation phase, we completed the quality control procedures that accompanied each of the earlier phases, which are described in further detail in the section below (Quality Control).

### Quality Control

Quality control was part of the harmonization process, applied first to the raw source data and then to the harmonized data. It comprised (i) quality control of the source data before harmonization, (ii) assessment of construct comparability for the multi-item psychosocial measures, (iii) quality control of the harmonization pipeline, and (iv) additional data quality control specific to downstream modeling, including the removal of implausible or out-of-range values. For (i), we manually examined each cohort and visit for variable ranges, missingness patterns, and the distributions of corresponding variables across cohorts and visits. Distributional differences flagged possible batch effects and differences in units or coding (for example, waist circumference recorded in centimeters in BPRHS and in inches in PROSPECT), which the harmonization transformations then resolved; in one case they showed that a feature with matching definitions could not be harmonized (see Construct Comparability). After harmonization, the quality control report produced by the pipeline (iii) checks a set of harmonized variables against their source variables and across cohorts, confirming that the transformations produced the expected results. The missingness and distributional summaries reported below are computed on the harmonized data.

Overall missingness differed by cohort and visit. Across all records in the harmonized dataset, 46.0% of values were missing in BPRHS and 59.1% in PROSPECT. Missingness was lowest at baseline in both cohorts (39.3% in BPRHS, 37.3% in PROSPECT) and higher at follow-up, reaching 53.7% at BPRHS visit 3 and 81.2% at PROSPECT visit 2. When stratified by visit and modality (Figure 1A), the highest values are for FFQ variables at PROSPECT visit 2 (95.9%), where the food frequency questionnaire was completed by only 103 participants, and for SDOH variables at BPRHS visit 4 (82.9%).

**Figure 1:**
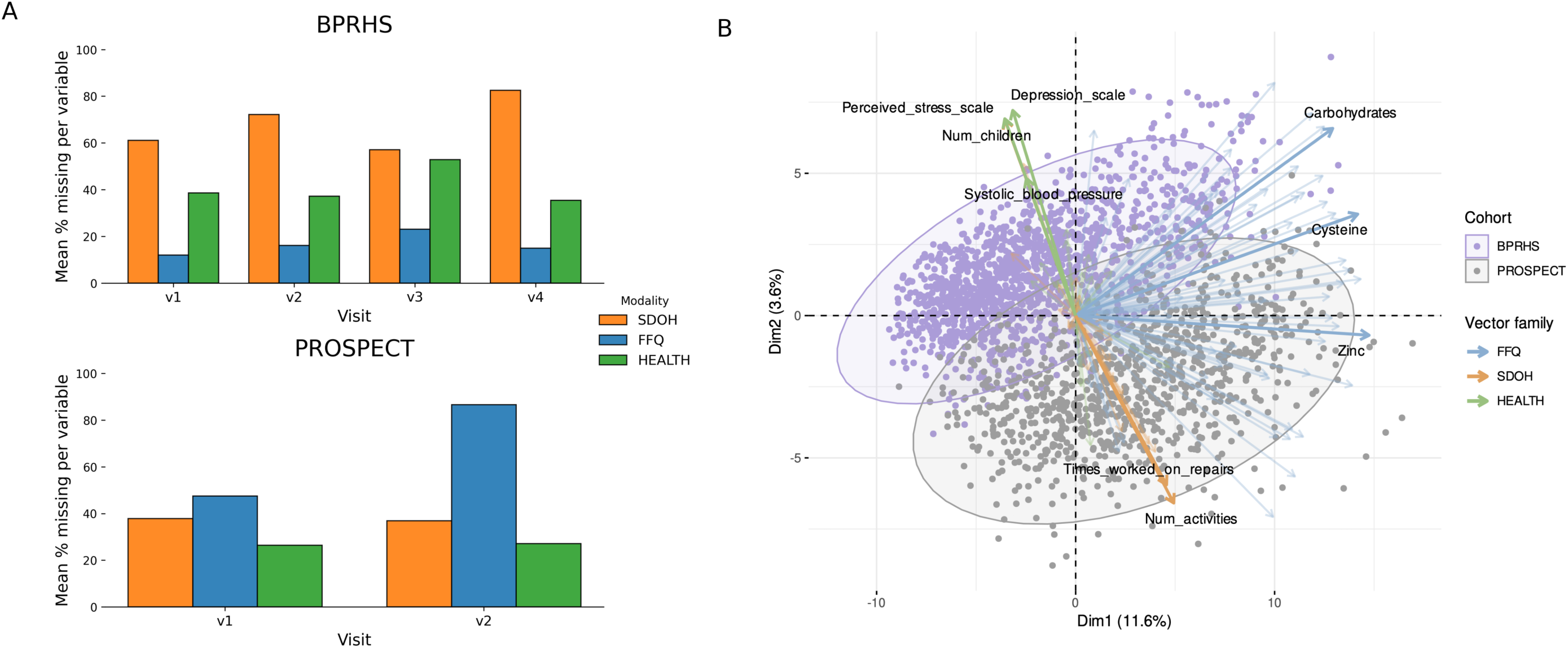
A) Missing data in the harmonized dataset by cohort, visit, and modality. Bars show the mean percentage of missing values per variable within each modality, computed over all records in the harmonized dataset for each cohort and visit; B) Multivariate structure of the integrated dataset showing cohort separation and variable loadings. Arrows show numeric loadings for FFQ, SDOH, and health variables, with darker arrows indicating variables with the largest loadings within each variable type.

We also examined observed correlates of missingness among harmonized variables. Logistic regression models were fitted for each numeric variable, predicting a binary missingness indicator (1 = missing, 0 = observed) from cohort, education, and perceived stress. These predictors were selected to capture structural (cohort), socioeconomic (education), and psychosocial (perceived stress) factors that could plausibly influence response completeness and data availability across harmonized variables. An association with an observed covariate indicates that missingness is not completely at random, but it does not distinguish MAR from MNAR, which cannot be determined from the observed data alone (19).

Across all numeric variables, missingness patterns were predominantly structured, with the majority (approximately 70-90%) showing an association with at least one of the tested predictors. For instance, missing waist circumference values were significantly associated with lower education and with the PROSPECT cohort, suggesting that data were missing due to systematic differences in study context rather than random omission.

These findings are consistent with domain- and cohort-specific missingness observed elsewhere in the dataset (e.g., higher FFQ missingness in PROSPECT and higher SDOH missingness in BPRHS), and indicate that missingness is driven largely by differences in survey coverage, instrument administration, and visit timing across cohorts.

Sources of missingness. Missingness in the integrated dataset arises from distinct processes, which we report separately because they have different implications for analysis (Table2, SI table 4). A value may be answered (code 0), not administered at that visit (code 1), absent because the participant missed that visit but returned later (code 2), unanswered by a participant who attended and was asked (code 3), not asked because of questionnaire routing (code 4), or absent because the participant was lost to follow-up (code 5). Across all participant-visits, 37.9% of values are answered, 31.9% are absent owing to loss to follow-up, 14.2% correspond to variables not administered at that visit, 3.9% were routed out by design, 0.4% belong to visits missed by participants who later returned, and 11.7% are item non-response.

The composition differs between cohorts and visits. In BPRHS, variables not administered account for 23.6% to 26.0% of values at visits 1 to 3, as instruments were added and withdrawn, item non-response ranges from 1.2% to 12.0% across visits, and loss to follow-up rises from 14.1% at visit 2 to 69.4% at visit 4. The highest item non-response, at visit 3 (12.0%), largely reflects 396 records that contain only FFQ data at that visit and whose identifiers could not be linked to a visit 3 clinic participant (Table 1), because these records have data at the visit, their empty SDOH and health variables are coded as unanswered. In PROSPECT, all harmonized variables were administered at baseline, but item non-response is higher (29.3%). At visit 2 the dominant process is loss to follow-up (69.7% of values), against 11.5% item non-response. The high overall missingness at PROSPECT visit 2 therefore reflects follow-up participation rather than data quality.

To describe distributional differences between cohorts, we computed the two-sample Kolmogorov--Smirnov (KS) test (20) for each harmonized numerical variable, which measures the maximum absolute difference between the empirical cumulative distribution functions (ECDFs) of the two samples. We report it as a descriptive diagnostic, given that cross-cohort differences may reflect genuine differences between mainland and island-based Puerto Rican populations, which the integrated dataset is intended to preserve, and distributional similarity does not establish that two measures capture the same construct.

The strongest cross-cohort differences were observed for social factors (such as household composition, social assistance, and social activities) and selected nutritional variables (Figure 2A and 2B), yet most variables exhibited moderate or low divergence of KS (approximately 87% with KS <0.3), indicating moderate or low distributional divergence.

**Figure 2:**
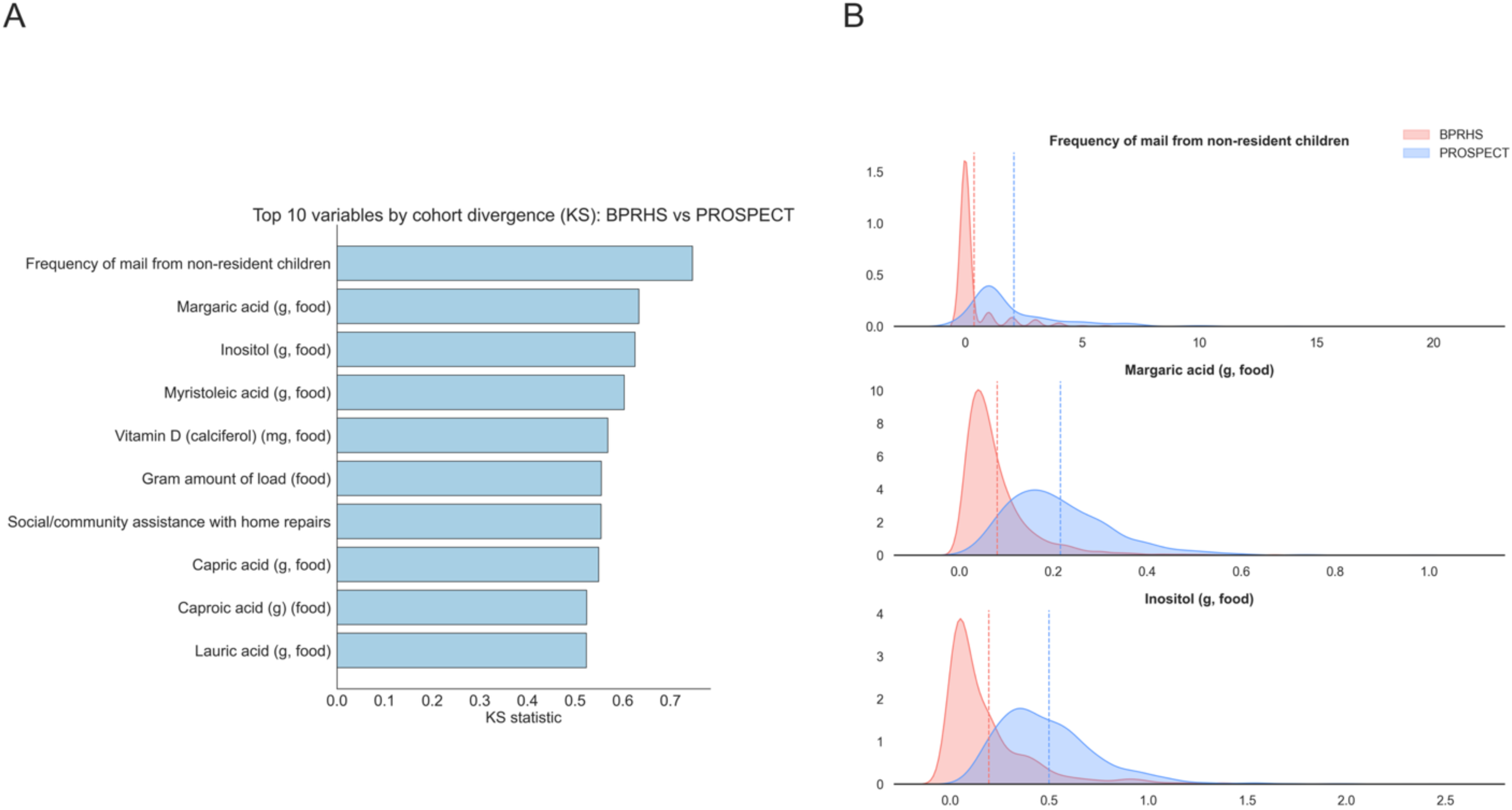
A) Top ten variables showing the largest distributional differences as per Kolmogorov--Smirnov (KS) statistics between the BPRHS and PROSPECT cohorts. Variables shown include: Frequency of mail from non-resident children (hmz_sdoh_soc5a), Margaric acid (g, food) (hmz_ffq_s17_0), Inositol (g, food) (hmz_ffq_inos), Myristoleic acid (g, food) (hmz_ffq_m14_1), Vitamin D (calciferol) (mg, food) (hmz_ffq_vd), Gram amount of load (food) (hmz_ffq_gramamt), Capric acid (g, food) (hmz_ffq_s10_0), Social/community assistance with home repairs (hmz_sdoh_soc_repairs), Caproic acid (hmz_ffq_s06_0) and Lauric acid (g, food) (hmz_ffq_s12_0). B) Kernel density estimates (KDEs) for the three variables with the largest KS divergence: Frequency of mail from non-resident children (hmz_sdoh_soc5a), Margaric acid (g, food) (hmz_ffq_s17_0), and Inositol (g, food) (hmz_ffq_inos).

These values indicate where the two cohorts diverge most, and identify variables whose harmonization decisions warrant closer inspection by users of the dataset before analysis.

For range inspections, we reviewed minimum and maximum values for all variables. Outliers or implausible entries were flagged for manual verification, and obvious data entry or coding errors were corrected when appropriate. These checks identified a small number of errors in the source data, including duplicated participant identifiers and implausible dates, which were resolved before harmonization and reported to the source institutions.

For (ii) the assessment of construct comparability of the multi-item psychosocial measures: We focused on three scales: the CES-D (20 items), the Perceived Stress Scale (14 items), and the Perceived Discrimination Questionnaire (8 items from section 1). We fitted multi-group confirmatory factor analyses with ordered-categorical indicators (WLSMV estimation with theta parameterization and identification constraints following (21) , comparing cohorts at baseline for the CES-D (complete-case n = 1,484 and 1,569) and the PSS (complete-case n = 1,480 and 1,604), and, for the discrimination items, the only BPRHS visit recording them (V3) against PROSPECT baseline (complete-case n = 946 and 1,594). Invariance was evaluated by successively constraining thresholds, then loadings, then intercepts to equality across cohorts, treating a step as non-invariant when Δ CFI < -0.010 and ΔRMSEA > 0.015 (22)Because item-level differences need not propagate to the summed scores, we also evaluated differential test functioning (23). For each scale we fitted a multiple-group generalized partial credit model, using the items that showed no appreciable strain in the factor analyses as anchors and freely estimating the remainder, and expressed the cohort difference in expected total score in raw points and as a percentage of the scale range.

All three scales met the criteria for scalar invariance (SI Table 5), in each case under the better-fitting factor structure (four-factor for the CES-D, two-factor for the PSS). Successive changes in CFI did not exceed 0.002 for the CES-D or the Perceived Discrimination Questionnaire, and reached 0.009 for the PSS at the final step - within the conventional threshold, but by a narrower margin. The residual strain is localized: the reverse-worded positive affect items of the CES-D, whose subscale reliability also differs across cohorts (0.59 and 0.75); two PSS items, one excluded from the shorter PSS-10; and, in the discrimination questionnaire, only the item on being criticized for one’s accent, plausibly reflecting the language of administration. Because analyses of the integrated dataset typically use summed scores rather than individual items, we also quantified differential test functioning (23). Expected total scores differed between cohorts by less than one point on every scale (signed 0.59 and unsigned 0.76 of 60 for the CES-D, and signed -0.46 and unsigned 0.54 of 56 for the PSS, and signed and unsigned 0.07 of 24 for the discrimination questionnaire). The item-level differences therefore do not materially affect the scores distributed in the harmonized dataset, supporting the inferential equivalence of the summed scores across cohorts, although these specific items warrant caution.

For (iii) quality control of the harmonization pipeline, we incorporated a validation step (Figure 3A) that produces a quality control report with three components: (A) checks of the dimensional consistency of the integrated dataset; (B) consistency between a set of harmonized variables, which may be extended by future users in the validate.py script provided in the harmonization pipeline and their corresponding raw variables, as well as consistency of the harmonized variables across BPRHS and PROSPECT; and (C) additional structural and content checks, including validation of expected variable presence and summaries of numeric versus non-numeric variables. This validation step allows future users to gain insight into the harmonization process and tailor their own validation procedures, ensuring flexibility and transparency in extending or adapting the pipeline for additional datasets or use cases.

**Figure 3:**
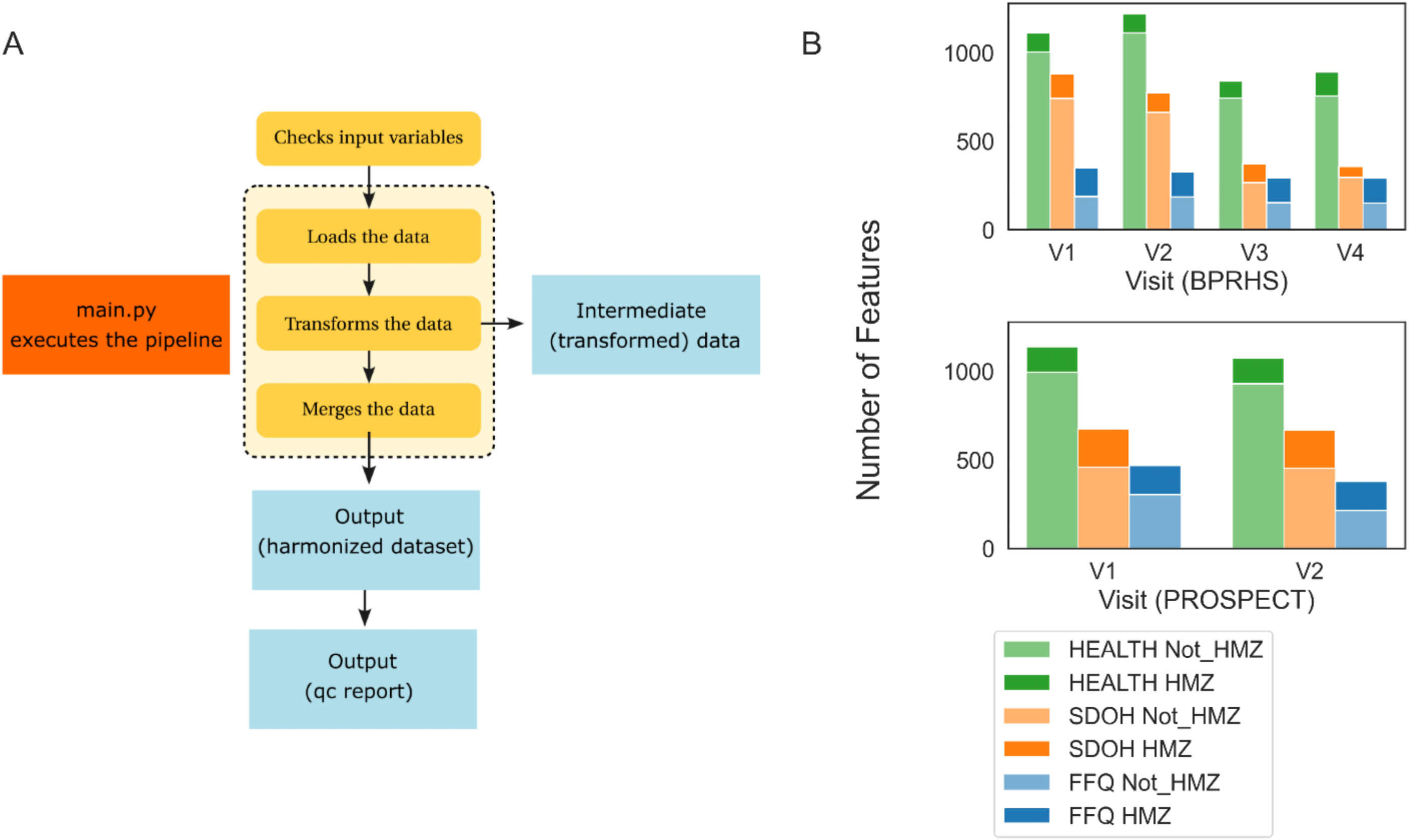
A) Schematic of Harmonization (HMZ) Pipeline; B) Features in the Not-Harmonized (Not HMZ) datasets that were used in this study, and in the Harmonized (HMZ) datasets, across different cohorts, visits, and modalities.

In addition to this internal validation step, we also make available a synthetic dataset, that was generated by sampling from normal distributions defined by the means and standard deviations of the original raw variables, which can be used to test the harmonization pipeline. The synthetic-data pipeline illustrates the full harmonization workflow and enables reproducible testing for users who may want to request access to the restricted raw data from the University of Massachusetts Lowell and Harvard University.

The evidence supporting comparability differs by class of variable: for syntactically harmonized features it follows from the variable definitions; for semantically harmonized features it rests on the decision rules documented in SI table 1; and for the multi-item psychosocial scales it was tested empirically (step ii).

In the (iv) quality control for the downstream RF model, we removed participants with implausible dietary energy intake values (<600 kcal or >4, 800 kcal), following established dietary assessment methods (16). We also excluded observations in which survey responses fell outside allowable codebook-defined categories (e.g., values outside a 1--4 response scale), as these represented data-entry errors. Prior to modeling, analyses were restricted to harmonized variables that were observed in both cohorts, exhibited less than 90% missingness within each cohort, and less than 50% missingness overall. In addition, participants with more than 10% missing values across the retained predictors were excluded. After filtering, the modeling dataset for Visit 1 included 1,407 BPRHS participants and 947 PROSPECT participants, with the smaller PROSPECT sample reflecting higher rates of missingness and implausible values. For the descriptive analyses of changes in diabetes status between visits, we applied a separate inclusion criterion focused on predictors required to calculate diabetes status. Specifically, we required non-missing diabetes status at both Visit 1 and Visit 2, yielding 1,258 BPRHS and 523 PROSPECT participants.

### Prediction of Diabetes Status: The Random Forest Model

Diabetes status was defined for each participant at the baseline (V1) and first follow-up (V2) for both cohorts using a definition of diabetes that incorporated laboratory and medication data. More specifically, participants were classified as having diabetes if glycated hemoglobin (HbA1c) was ≥6.5% (24), fasting glucose was ≥126 mg/dL, or diabetes medication use was reported. This binary outcome was calculated for the first two visits for each cohort, since PROSPECT only included two visits. All predictors were drawn from V1, and models were trained to predict diabetes status concurrently at V1 and prospectively at V2. To prevent target leakage, the variables used to define the outcome (HbA1c, fasting glucose, self-reported diabetes diagnosis, age at diagnosis, current diabetes status, and diabetes medication use), together with serum insulin, participant identifiers, visit, and days since visit 1, were excluded from the predictor set.

Random Forest (RF) classifiers were trained to predict diabetes status using the tidymodels framework in R. RFs employ an ensemble of decision trees and are robust to nonlinear and non-monotonic relationships as well as complex interactions among predictors (25). Pre-processing was implemented using the recipes framework in R and comprised imputation of missing values, removal of near-zero variance features, normalization of continuous features, and correlation filtering. Because the RF workflow requires complete predictor data, missing values were imputed using k-nearest-neighbors imputation for numeric features and mode imputation for categorical features. Imputation was estimated on the training data of each split and applied unchanged to the corresponding test data, so that no information from the test set entered the imputation. We used single rather than multiple imputation because the aim of the worked example is prediction rather than unbiased estimation of population parameters and their standard errors, and k-nearest-neighbors imputation draws on the observed values of similar participants and therefore retains the multivariable structure of the predictors. These methods assume that data are missing at random, which cannot be verified from the observed data. To assess sensitivity to the imputation approach, we repeated the pooled V1 analysis with median imputation in place of k-nearest-neighbors imputation, and with missingness-indicator features added so that the model could use informative missingness (SI Table 2). These analyses do not test robustness to data missing not at random. Imputation was applied only within the RF workflow; the integrated dataset contains no imputed values. Feature selection was conducted with the Boruta algorithm (26), retaining only features classified as confirmed.

**Table 2:**
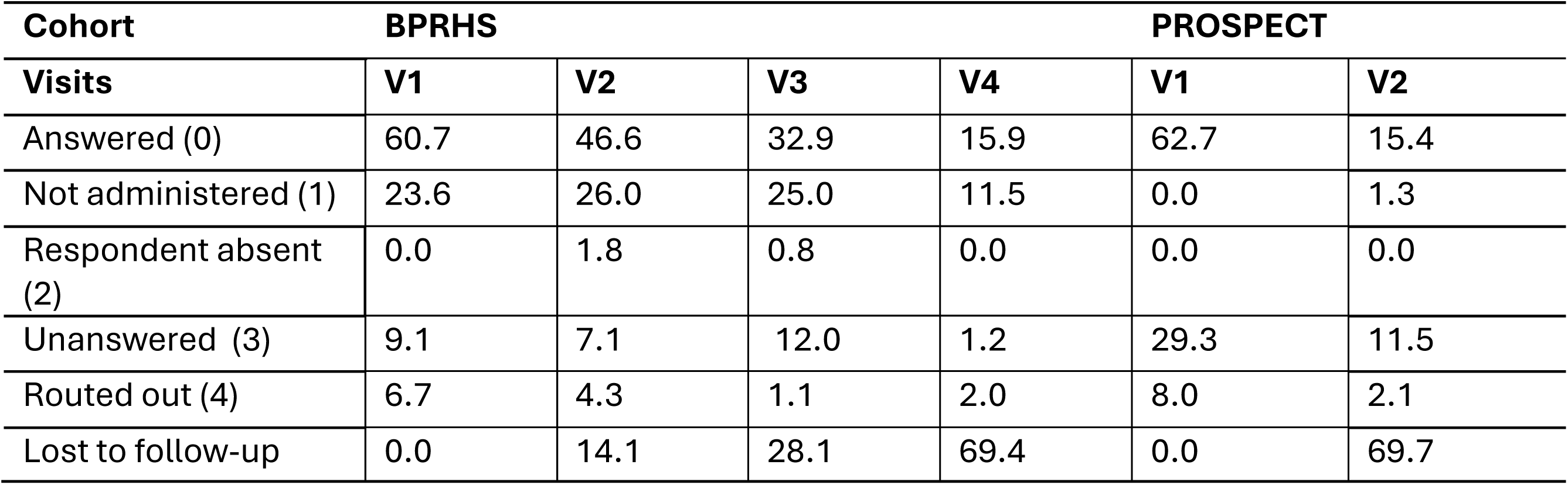
Sources of missingness in the harmonized dataset, by cohort and visit. Values are the percentage of all cells of harmonized variables for participants in the given cohort and visit. Participants are included from their first visit with data onwards, including visits they missed.

The workflow was repeated over ten random 75/25 training–test splits stratified by diabetes status, with cohort- and outcome-specific case weights to account for unequal cohort sizes and class imbalance, and hyperparameters tuned by five-fold cross-validation within each training set (details in SI Methods). To describe how well models generalize across cohorts, we compared four training regimes: within each cohort, pooled, pooled training with testing on one cohort, and training on one cohort with testing on the other.

## Results

We report two main outcomes. The first is the harmonized BPRHS/PROSPECT resource: the integrated dataset, the documented pipeline that produces it, and metadata on variable definitions, transformations, availability, and missingness across cohorts and time points. The second is a worked example of a downstream analysis using this resource, comprising descriptive patterns of diabetes status across visits and a Random Forest model predicting diabetes status.

### Harmonized BPRHS/PROSPECT Resource

#### Integrated Dataset

The integrated dataset features a total of 539 features (208 SDOH features, 167 Health features, and 164 FFQ features (Table 3). As shown in Figure 3B, the integrated dataset is a relatively small subset of the original data from the BPRHS and PROSPECT studies, but it is more balanced across modalities and visits, when compared with the original data that we used in this research. As mentioned earlier, all harmonized features were renamed and added prefixes, standing for harmonization, as well as the different modalities of data, and groups of features within each modality. Among the harmonized data, 375 features underwent some additional transformation, aside from renaming, to ensure comparability and compatibility (SI Table 1).

**Table 3:** Overview of groups of features in modalities SDOH, HEALTH and NUTRITION in the integrated dataset (more detailed description of variables is provided in the metadata SI Table 1).

| **SDOH** | **HEALTH** | **NUTRITION** |
| --- | --- | --- |
| Demographic and Education* | Laboratory | Diet Type |
| Household Composition | Anthropometrics | Meal Place |
| Neighborhood Food and Activity | Medical Diagnoses | Supplements |
| Work History and Income | Family History | Nutrient intakes |
| Social and Community Support and Assistance | Health Behaviors |  |
| Access to Healthcare | Perceived Stress |  |
| Perceived Discrimination | Depression |  |
| Total SDOH: 208 | Total HEALTH: 167 | Total NUTRITION: 164 |
| Total: 539 |  |  |

*\*For our analysis, Age and Sex were considered both SDOH and HEALTH features.*

We examined the multivariable relationships among cohorts and variable modalities in the integrated dataset using factorial analysis of mixed data (FAMD) (27) (Figure 1B). In the space defined by the first two FAMD dimensions, individuals from the two cohorts formed partially overlapping clusters. The BPRHS cluster lay in the direction of higher depressive symptom scores, higher perceived stress, a greater number of children, and higher systolic blood pressure. Several health-related variables loaded together along a common vertical axis (Dimension 2). In contrast, several SDOH variables, including the frequency of working on home repairs and the number of social or community activities, loaded in the opposite direction along this same axis and were oriented toward the PROSPECT cluster, consistent with higher values for these SDOH measures among PROSPECT participants. Dietary variables from the FFQ defined a second, largely orthogonal axis relative to this health-SDOH gradient. Higher intakes of carbohydrate, cysteine, and zinc were among the strongest-loading dietary variables and loaded together along this horizontal dimension, reflecting correlated dietary patterns that are distinct from the health-related dimension.

These patterns indicate that the harmonized variables retain coherent and interpretable multivariate structure, and that the integrated dataset supports joint analysis across the two cohorts. However, they do not establish measurement equivalence. Because the cohorts differ in age, geographic setting, socioeconomic circumstances, and health, the degree of overlap between them in this space reflects both harmonization and genuine population differences.

#### Harmonization Pipeline

The harmonization pipeline (Figure 3A) includes five modular steps (i) *Check*: feature check to confirm the presence of a core set of features in the input datasets, essential to the harmonization process, thus allowing users of the pipeline working with slightly different versions of the datasets used in this harmonization; (ii) *Loading*: Data were loaded in three different modalities, Health, SDOH and FFQ); (iii) *Transform*: Data were transformed to enable harmonization of features across visits and cohorts; (iv) *Merge*: The transformed Health, SDOH, and FFQ data were merged to produce the final BPRHS/PROSPECT harmonized and integrated dataset; and (v) *Validation*: We included a validation and quality control check, which assesses structural quality control (dimensions, duplicates, shape, column and row counts, cross-domain overlap, high missingness columns) and a variable-level check comparing harmonized variables with their corresponding raw variables). The individual variables to be inspected were specified as a simple extensible lists of pairings (cohort, visit, raw variable, harmonized variable). The validation report is documented in a report that is one of the outputs of the harmonization pipeline. The harmonization pipeline is publicly available at https://github.com/rbatorsky/diabetes-cohort-harmonization.git.

Access to the original BPRHS and PROSPECT data requires submitting a data request through the established protocols at UMass Lowell and Harvard University, which ensures compliance with each study’s governance and participant protections.

### Worked Example: Predicting Diabetes Status

#### Diabetes Status Across Visits

When examining changes in diabetes status between visit 1 and visit 2 (Figure 4A), we restricted the analysis to participants with non-missing diabetes classifications at both timepoints (BPRHS: n=1,258; PROSPECT: n=523). In both cohorts, most participants maintained the same diabetes status across visits (BPRHS: 1,063 individuals, 84.5%; PROSPECT: 487 individuals, 93.1%). Changes in status were more common in BPRHS, where 102 participants (8.1%) showed progression and 93 participants (7.4%) showed remission. In PROSPECT, changes were less frequent, with 19 participants (3.6%) progressing and 17 participants (3.3%) remitting. A multinomial logistic regression adjusting for age and time between visits showed that cohort membership was a statistically significant predictor of both remission and progression (p=0.002 and p=0.005, respectively), with PROSPECT participants having significantly lower odds of changing diabetes status in either direction compared with BPRHS. Because changes in status were rare, particularly in PROSPECT (36 of 523 participants), the worked example predicts diabetes status at each visit rather than change between visits; exploratory models of progression performed close to the prevalence baseline.

**Figure 4:**
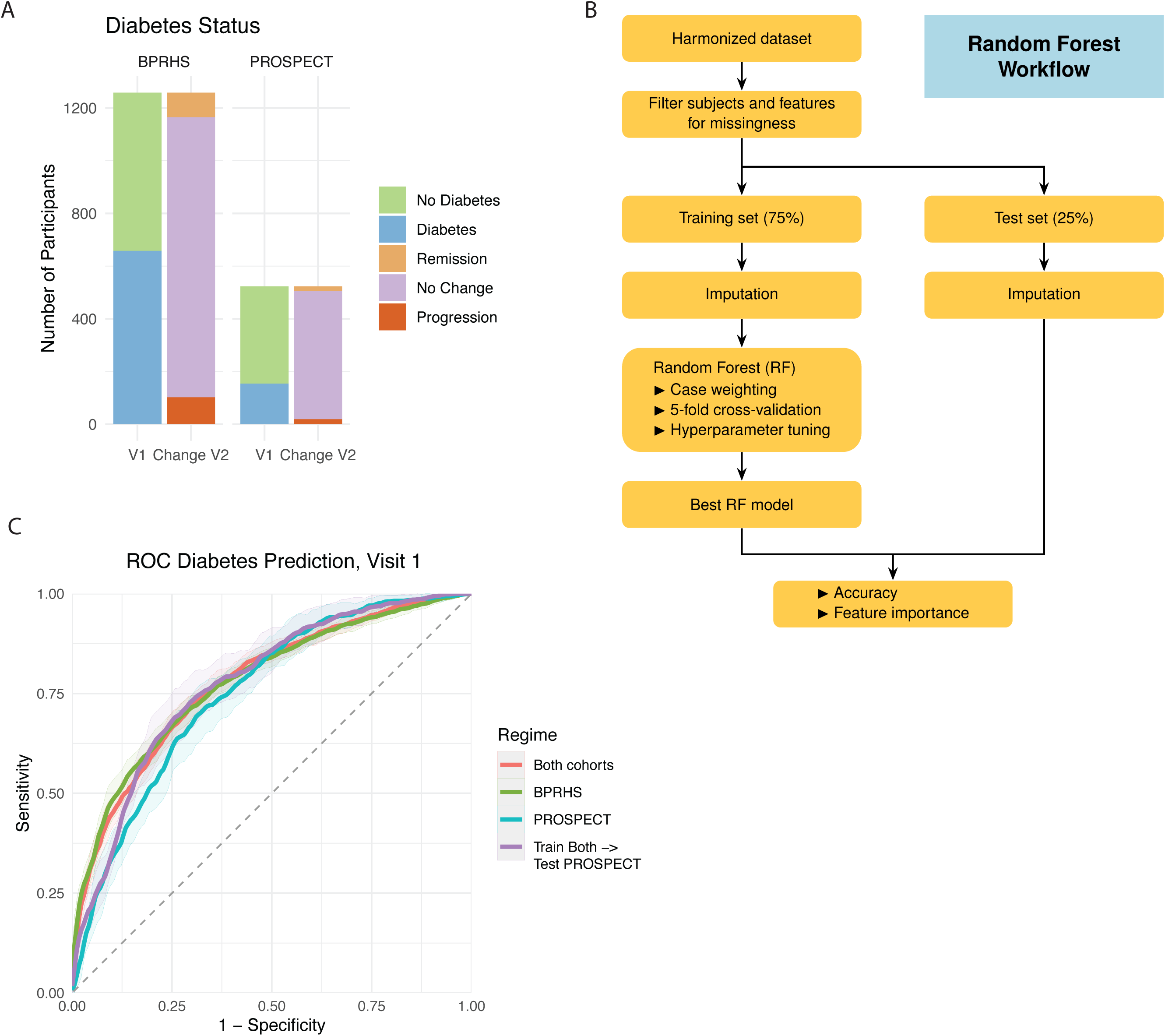
Random Forest model to predict diabetes status from harmonized visit 1 data A) Diabetes status and status change in visit 1 and visit 2 in both BPRHS and PROSPECT cohorts B) Schematic of the RF modeling workflow C) ROC curves across 10 random train–test splits for each modeling regime, illustrating within-cohort and cross-cohort discriminability.

#### Random Forest Model

We used RF models (Figure 4B) to predict diabetes status concurrently at V1 and prospectively at V2. This worked example was intended to illustrate a downstream use of the integrated dataset and to evaluate whether learning from pooled cohorts could improve predictive performance.

The ROC curve for the model trained on the harmonized dataset closely tracked that of the BPRHS-only model, the cohort with more participants in the modeling dataset (Figure 4C).

The ROC curves of the different regimes crossed, indicating that the relative discriminability of each cohort depends on the operating point. At high specificity, PROSPECT appears more difficult to classify. At moderate false-positive rates, however, the pooled-training model tested on PROSPECT performed best of all regimes. This suggests that the additional BPRHS training data helped the model recover signal in the smaller cohort (SI Results).

AUC was similar for the pooled and BPRHS-only models (0.778 and 0.779), and training on one cohort and testing on the other gave slightly lower values (0.758–0.762). PROSPECT benefited most from pooling: training on both cohorts raised its AUC from 0.752 (PROSPECT-only) to 0.776, close to the BPRHS value. Accuracy must be compared in the context of differing prevalence of diabetes in PROSPECT than in BPRHS (28% vs 53%). A model predicting no diabetes for every PROSPECT participant would already reach 72% accuracy, and only the pooled-training model clearly exceeded this (0.747, vs 0.717 for PROSPECT-only, which did no better than this baseline). Discrimination was slightly lower at V2 (pooled AUC 0.748), as expected when V1 features are used to predict status at a later visit. Taken together, the harmonized feature set retained its ability to discriminate diabetes status across cohorts, and pooling improved prediction in the smaller cohort without reducing it in the larger one.

Permutation importance (Figure 5A) ranked health and dietary features highest. In the pooled and BPRHS-only models, aspartame intake was the most important feature, followed by lipid and adiposity markers (plasma triglycerides, LDL, waist circumference, systolic blood pressure and BMI). The PROSPECT-only model was dominated by clinical and anthropometric variables, and aspartame ranked much lower. Cardiometabolic biomarkers were therefore central in every model, whereas the importance of dietary features varied by cohort (SI Results).

**Figure 5:**
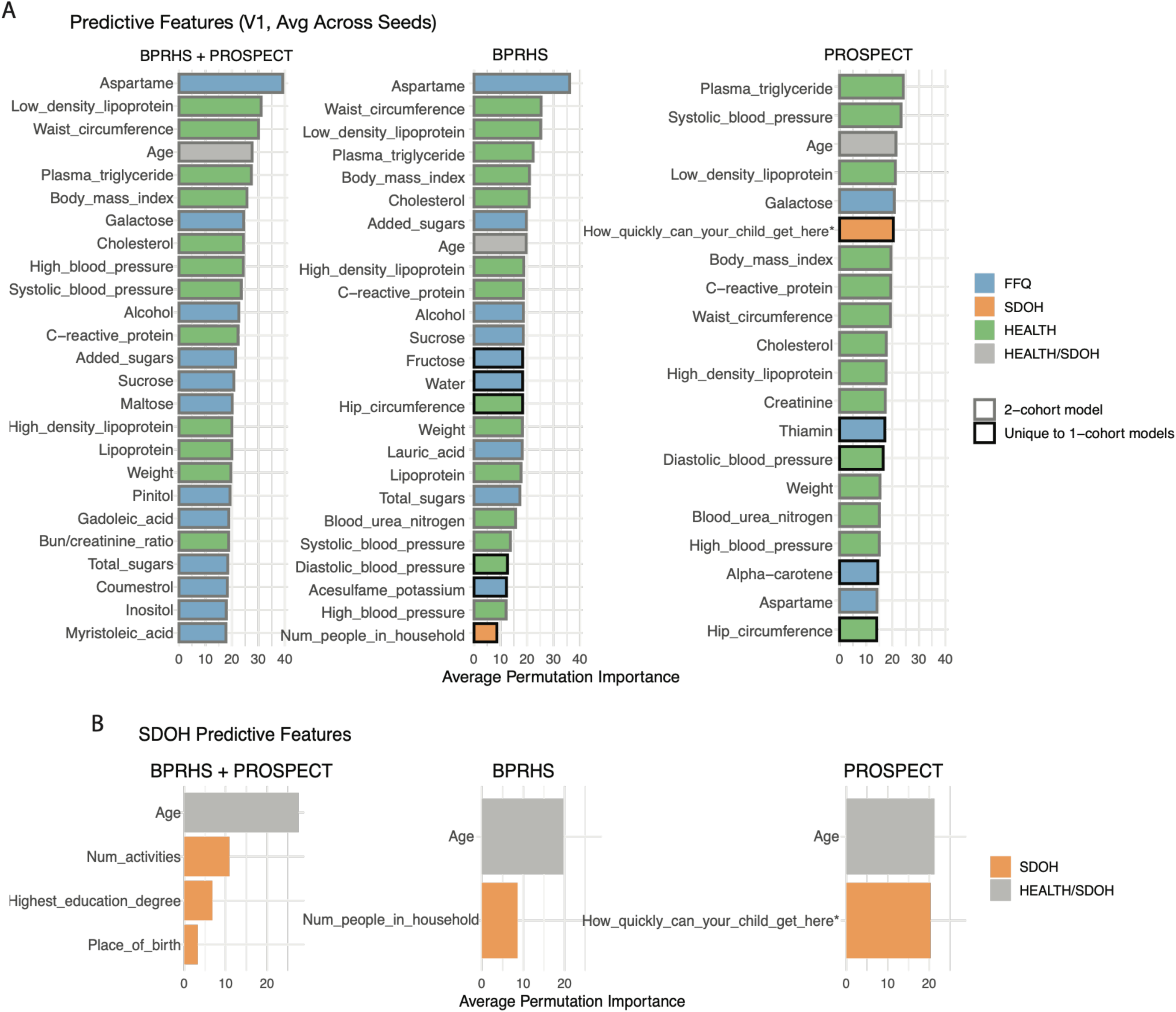
Permutation feature importance for RF models predicting diabetes status at V1 A) Top-ranked features across three models trained on harmonized (BPRHS + PROSPECT), BPRHS-only, and PROSPECT-only data. Features are colored by data modality and ordered by mean permutation importance in the harmonized model; only features retained by the Boruta algorithm are shown B) Importance of SDOH features only.

SDOH features contributed less than health and dietary features, apart from age, which is treated as both a health and an SDOH feature. Among SDOH features (Figure 5B), the pooled model ranked number of social activities, highest education degree and place of birth highest, the BPRHS-only model the number of people in the household, and the PROSPECT-only model the time a non-cohabiting child would take to reach the respondent in case of need. The distributions of the top-ranked features by cohort and diabetes status are shown in SI Figures 1 and 2.

## Discussion

This study shows how two independently collected cohorts can be integrated into a single documented resource, with the basis for each variable’s comparability recorded, so that they can be analyzed jointly. As a worked example, a Random Forest model trained on the integrated data performed as well as the BPRHS-only model, improved classification in PROSPECT, and identified cardiometabolic predictors that were consistent across cohorts, whereas the importance of dietary features varied, illustrating the kind of analysis the resource supports.

The open-source pipeline and accompanying structured metadata further support transparency and reuse by enabling other researchers to reproduce the integration process or adapt it to similar cohort studies. In this context, transparency functions not only as documentation but as a mechanism for empirical assessment of integrated data resources.

This case study provides a practical example of how documented integration and empirical evaluation of predictive performance can support secondary use of complex longitudinal cohort data, particularly in population studies collected under different protocols.

### Diabetes Status Across Visits

Diabetes status was stable between V1 and V2 for most participants in both cohorts, even allowing for the different timing of the visits. Changes were more common in BPRHS, where progressions (no diabetes at V1, diabetes at V2) were slightly more frequent than remissions (diabetes at V1, no diabetes at V2) (8.1% vs 7.4%). Progressions more plausibly reflect incident type 2 diabetes with aging and accumulating metabolic risk. Remission in later life is less well characterized; normal glucose regulation can be restored through preserved insulin secretion, weight loss, healthy eating and physical activity (28, 29), but in our data remissions more likely reflect borderline cases that fell below the diagnostic thresholds with treatment or lifestyle change, or measurement variability. These interpretations remain hypotheses.

### Predictive Performance of the Integrated Dataset

The RF model that we developed for the classification of diabetes status does not identify causality, but rather predictive associations. The model trained on the harmonized data compares well with other models reported in the literature, with predictive accuracy of approximately 70% (30, 31). The model showed similar performance with the model trained in the BPRHS data, and both outperformed the models trained in PROSPECT data. This pattern shows that the integrated dataset supports diabetes classification at a level comparable to that of the larger single cohort. At the same time, the relative underperformance of the model trained on PROSPECT data, likely reflects cohort-specific factors, such as smaller sample size.

Our results indicate that features showing higher predictive power for diabetes in the harmonized cohort are related to nutrition and health. In both the model trained in harmonized data and BPRHS data, aspartame, an artificial sweetener that provides sweetness without adding calories, is the top predictor of diabetes status. Importantly, aspartame should be interpreted as a marker rather than a direct causal effect; its prominence in the model most plausibly reflects a common clinical recommendation for individuals diagnosed with diabetes to substitute aspartame for caloric sweeteners as part of dietary management (32).

The clustering of age, adiposity, lipids, and blood pressure at the top of the permutation-importance profiles is consistent with established markers of T2D risk. Advancing age reflects greater cumulative metabolic exposure (33, 34); excess adiposity (BMI, waist circumference) promotes insulin resistance (35); dyslipidemia (triglycerides, LDL) tracks hepatic insulin resistance (36, 37); and elevated blood pressure (systolic pressure, hypertension) signals vascular dysregulation that commonly accompanies the insulin-resistant phenotype (38, 39).

Dietary features, including sugars and fatty acids, generally ranked below health-related markers but consistently appeared among the higher-importance features in the pooled and BPRHS models, and less so in PROSPECT, which may reflect cohort differences in intake distributions or exposure measurement. As with aspartame, permutation importance reflects predictive value rather than causality, and these features may be proxies for broader eating patterns or for dietary changes following medical advice.

In our models, SDOH features contributed less to prediction than health and dietary features. In the pooled model, the most informative were the number of social activities, highest education degree and place of birth (Figure 5B, SI Figure 2), which may serve as proxies for income, a known risk factor for diabetes (40–44), or reflect overall health and social connection.

### Limitations of the Integrated Dataset

Integrating independently collected cohort data necessarily involves trade-offs between comparability and detail. A well-documented concern is the harmonization–information trade-off, whereby increasing comparability across studies may reduce the level of detail retained in the integrated data (45). The magnitude of this trade-off depends on factors such as the availability and relevance of source data, the quality and transparency of data collection procedures, and the underlying comparability of the constructs being aligned (45).

In this study, we performed granular feature-level harmonization. While this approach improved consistency and comparability across datasets, it inevitably introduced information loss and reduced conceptual specificity (6). Certain nuances in the original variables, including culturally specific response categories, had to be generalized or omitted to enable alignment. As mentioned earlier, only a subset of available variables were harmonized (Figure 3B). Nevertheless, the number of features harmonized across visits and cohorts remained relatively consistent, reflecting an effort to preserve core content across modalities and timepoints while maintaining comparability.

Another limitation concerns the generalizability of cross-cohort comparisons. Non-substantive differences across studies, such as sampling strategies, timing of follow-up interviews, question phrasing, recall timeframes, survey flow, and language (10), can influence participant responses and complicate cross-study comparisons. We addressed these challenges by applying consistent feature-level transformations, aligning response categories, and standardizing derived variables across cohorts and visits. Nevertheless, residual methodological differences, particularly in survey-based measures, may persist and should be considered when interpreting harmonized results.

The interpretability of integrated survey data depends on both the quality of the original measurements and the alignment procedures used during integration (14, 45). To support comparability, we conducted a detailed review of documentation and metadata when aligning features across cohorts. Nevertheless, cultural differences reflected in question wording and response options, as well as variation in clinical examination procedures (e.g., anthropometry and blood pressure) conducted at different sites and time periods, may still influence the resulting variables.

We also acknowledge that integration cannot fully eliminate differences in measurement practices and interpretation across studies. Although our approach emphasizes transparency and empirical evaluation through downstream analysis, the reliability of integrated survey and examination variables ultimately depends on the quality, consistency, and comparability of the original documentation, metadata, and measurement protocols across cohorts (14, 45). Despite detailed codebook review and empirical evaluation, residual differences related to cultural context, question interpretation, and site- and time-specific measurement practices may remain. This is particularly relevant for clinical measures such as anthropometry and blood pressure, which were collected across different locations and years using potentially heterogeneous protocols.

Looking forward, integration of diverse population datasets such as BPRHS and PROSPECT could evolve toward multi-tier data architectures that preserve both raw and derived representations, enabling analyses at different levels of abstraction. Coupling transparent integration with ontology-based metadata (e.g., PhenX) and automated provenance tracking may further improve reusability and cross-study interoperability.

## Conclusions

In this study, we developed and documented an integrated dataset combining two longitudinal population cohorts related to Puerto Rican communities (BPRHS and PROSPECT). We implemented an open-source Python integration pipeline together with structured metadata describing variable definitions, source questions, transformations, and availability across cohorts and timepoints to enable transparency and reproducibility, as well as an explicit framework for establishing comparability between variables, distinguishing syntactic from semantic harmonization and reporting the empirical evidence supporting each class of feature. This documented approach illustrates how heterogeneous longitudinal data collected under different study designs and measurement protocols can be combined while retaining interpretable structure suitable for secondary population health research.

As a worked example of downstream use, we conducted a Random Forest analysis predicting diabetes status. The model trained on the integrated dataset matched the performance of the BPRHS-only model, and cross-cohort training modestly improved discrimination in PROSPECT relative to training on PROSPECT alone, illustrating that pooling cohorts can benefit the smaller dataset without sacrificing performance in the larger one. The cardiometabolic features driving prediction were consistent across cohorts, dietary features ranked highly in the pooled and BPRHS models, and SDOH features, while contributing less overall discriminative power, provided contextual signal relevant to population health interpretation. This example shows what an end-to-end analysis of the integrated dataset requires and what it can support, from variable selection through model specification to evaluation.

Overall, these findings illustrate how transparent integration, documented harmonization decisions, and evidence of construct comparability can support secondary analysis of complex longitudinal population data. More broadly, documenting integration and examining analytical behavior may inform future cross-cohort integration of similar population studies in which related populations are studied under different contexts.

### Data & Code Availability

#### BPRHS Datasets & Codebooks

The BPRHS datasets used in the harmonization pipeline can be requested through the University of Massachusetts Lowell (UMass Lowell) by submitting a CPH Analysis and Data Request Proposal Form, which is available at https://www.uml.edu/research/uml-cph/research/BPRHS/. The corresponding BPRHS codebooks are available at https://www.uml.edu/research/uml-cph/research/BPRHS/.

#### PROSPECT Datasets & Codebooks

The PROSPECT datasets used in the harmonization pipeline can be requested through Harvard University by submitting a Proposal Form, which is available at https://hsph.harvard.edu/research/puerto-rico-PROSPECT/data-requests/.

#### Harmonized Data & Metadata

To generate the integrated dataset, the input BPRHS and PROSPECT datasets must be provided to the harmonization pipeline implemented in Python. The harmonization code is available on GitHub (https://github.com/rbatorsky/diabetes-cohort-harmonization.git), and execution of the pipeline produces the integrated dataset (.csv file). The corresponding metadata is also available through GitHub. To test the harmonization pipeline, synthetic datasets are also provided in the Github repository, but original BPRHS and PROSPECT data must be requested from UMass Lowell and Harvard University as explained above.

#### Code Availability

The harmonization and modeling codes required to reproduce the results presented in this manuscript are available on GitHub (https://github.com/rbatorsky/diabetes-cohort-harmonization.git). Additional details regarding the code are provided in the ReadMe file, also available on GitHub.

## Author Contributions

Andreia Martinho provided research direction, developed the harmonization code and pipeline, produced visualizations, and co-wrote the manuscript. Rebecca Batorsky provided research direction, developed the data quality control and modeling pipeline, produced visualizations and co-wrote the manuscript. Martha Tamez, Josiemer Mattei, and Elizabeth Petit provided support on the PROSPECT dataset and feedback on the manuscript. Sabrina E. Noel and Katherine Tucker provided support on the BPRHS dataset and feedback on the manuscript. Bayan Tuffaha made contributions for the modeling. Jose Ordovas and Abani Patra provided feedback on the research direction and manuscript.

## Data Availability

All data produced in the present study are available upon reasonable request to the authors.

## Acknowledgments

We acknowledge the funding agencies that supported this research, including the National Institute on Minority Health and Health Disparities (NIMHD), National Institutes of Health (NIH), and National Institute on Aging under awards P01AG023395, P50HL105185, R01MD016584, R01HL143792, K01HL120951, and R21MD013650. We acknowledge the participants in the BPRHS and PROSPECT studies. We also acknowledge the author collaborators involved in BPRHS and PROSPECT data acquisition and codebook development, as well as Ziqi Zhou for initial exploratory data analysis during the research internship. Finally, we acknowledge the Tufts University High Performance Compute Cluster (https://it.tufts.edu/high-performance-computing), which was utilized in this research.

## Supplementary Appendices

SI Material. Full specification of the Random Forest workflow: pre-processing, feature selection, training–test splits, case weighting, hyperparameter tuning, and the four cross-cohort training regimes (SI Methods). Additional Random Forest results: ROC curves by training regime, feature importance beyond the leading predictors, distributions of top-ranked predictors by cohort and diabetes status, and sensitivity of the results to the imputation approach (SI Results).

**SI Figure 1.**
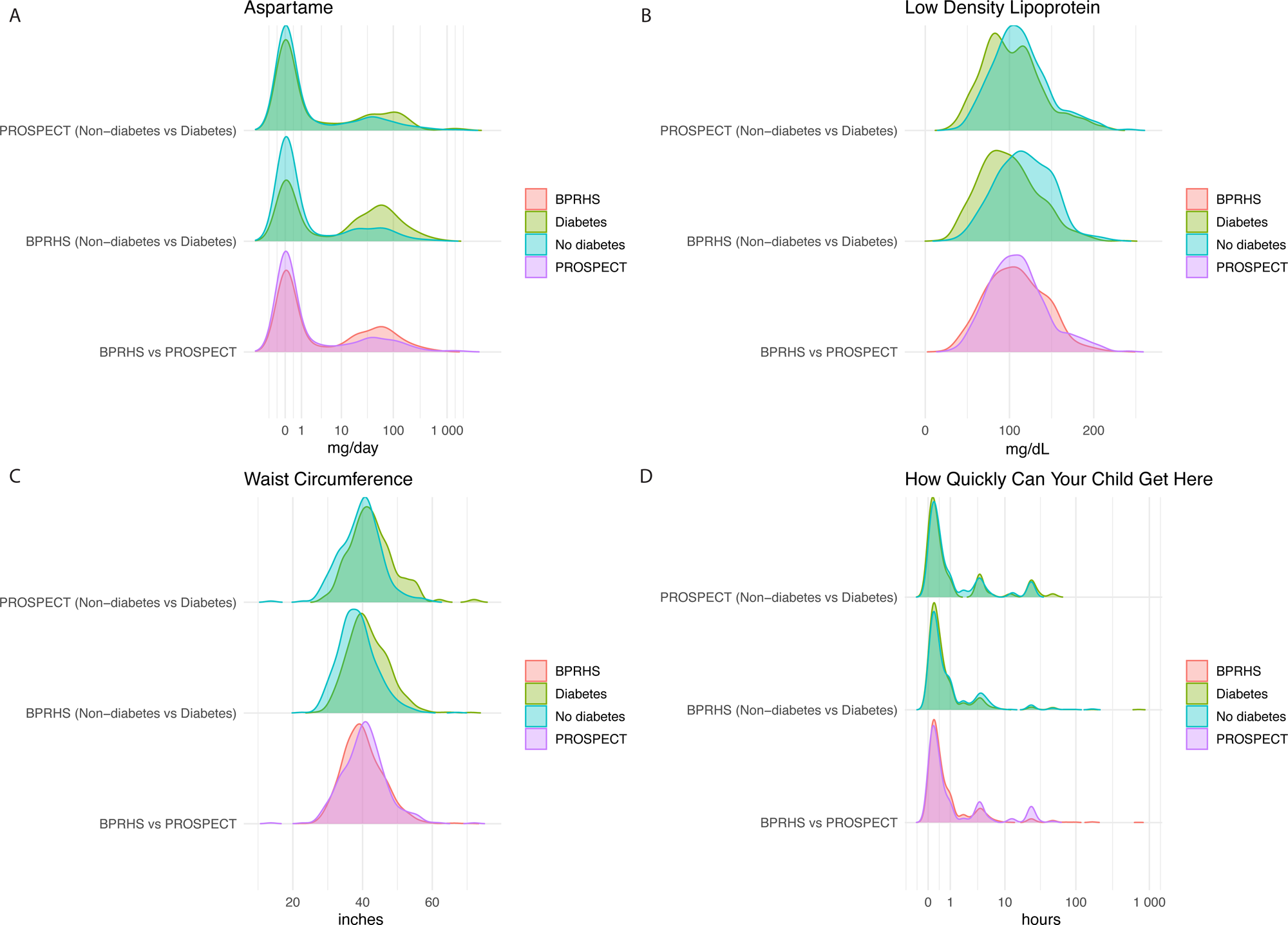
Distributions of selected predictive features from the pooled Random Forest model, by cohort and diabetes status.

**SI Figure 2.**
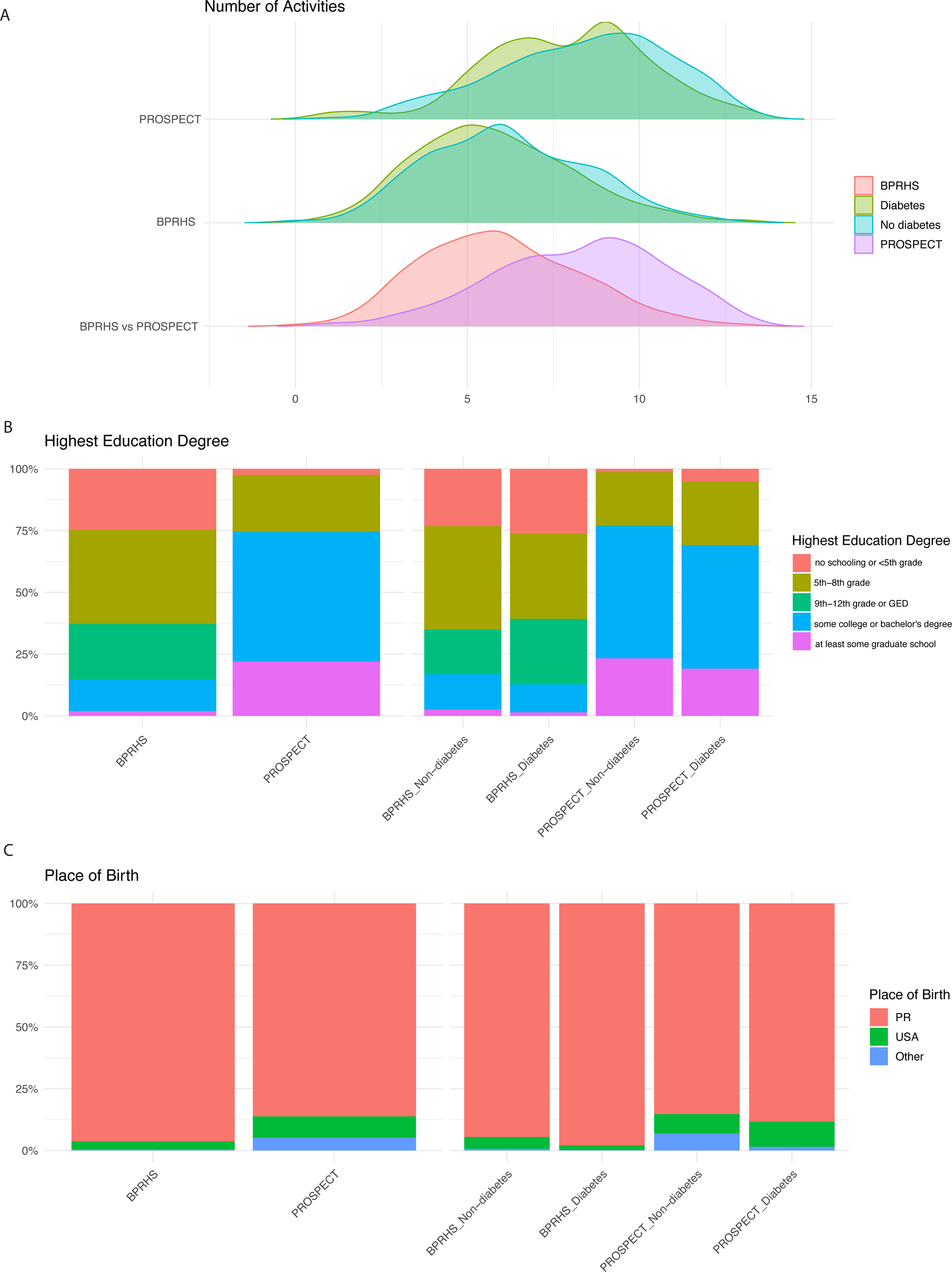
Distributions of the top-ranked SDOH features from the pooled Random Forest model, by cohort and diabetes status.

SI Table 1. Metadata of the harmonized dataset. For each variable: modality, type, definition, the transformations applied, whether it is available at each cohort and visit, and, for each visit, the source variable, its original question wording and response categories, and its source file. Additional sheets list the source files and define each column of the metadata.

SI Table 2. Sensitivity of the Random Forest worked example to the imputation approach. Test AUC, accuracy, and feature overlap for the pooled V1 model under k-nearest-neighbors imputation, median imputation, and k-nearest-neighbors imputation with missingness indicators.

SI Table 3. Cohort and visit characteristics relevant to harmonization. For each cohort and visit, including the separate FFQ waves: calendar years of data collection, interval between visits, languages of administration, instruments administered, changes in instruments between visits, recall periods, examples of differences in question wording and response categories, and differences in clinical measurement protocols.

SI Table 4. Cell-level missingness codes. One record per participant per visit, from the participant’s first visit with data onwards. Each harmonized variable is coded 0 (answered), 1 (not administered), 2 (respondent absent), 3 (unanswered), 4 (routed out) or 5 (lost to follow-up), as defined in Table 2.

SI Table 5. Measurement invariance and differential test functioning of the CES-D, Perceived Stress Scale and Perceived Discrimination Questionnaire across cohorts. (A) Single-group fit of alternative factor structures, by cohort; (B) measurement invariance sequence for the retained structure; (C) differential test functioning of summed scores; (D) composite reliability of scales and subscales, by cohort.

## AI Disclosure Statement

The authors used Claude (Claude Opus 5.5, Anthropic) for language editing, code generation and debugging. The output was reviewed and verified by the authors.

^1^ Variable_modality indicates the data modality of each harmonized feature (SDOH, FFQ, HEALTH). “Administrative” denotes variables identifying participant, cohort and visit, which do not belong to a data modality and are excluded from modality-level summaries.

