## Supplementary material for "Integrating Longitudinal Cohorts for Population Health Research: Mainland and Island-based Puerto Rican Diabetes Study": SI material

### SI Methods: Random Forest workflow

Random Forest (RF) classifiers were trained to predict diabetes status using the tidymodels framework in R. RFs employ an ensemble of decision trees and are robust to nonlinear and non-monotonic relationships as well as complex interactions among predictors (1).Pre-processing was implemented using the recipes framework in R, including mode imputation for categorical features, k-nearest-neighbors imputation for numeric features, removal of near-zero variance features, normalization of continuous features, and correlation filtering. Feature selection was conducted with the Boruta algorithm (2), retaining only features classified as confirmed, meaning their importance was consistently greater than randomly permuted features.

We generated ten independent random data splits, each stratified by diabetes status into 75% training and 25% testing sets, and repeated the entire workflow, from pre-processing through feature selection, hyperparameter tuning, and model fitting, for each split. To address unequal dataset sizes and class imbalance while retaining all available data, we used a case-weighting approach. In each split, cohort- and outcome-specific weights were computed as the inverse of each class frequency within BPRHS and PROSPECT, normalized to have mean one. These weights were then supplied to the RF during training via tidymodels::add_case_weights() functionality, ensuring that minority classes within each cohort contributed proportionally during model fitting. This approach mitigated imbalance without discarding observations. The held-out test sets were left unchanged to preserve natural prevalence. Hyperparameters mtry and min_n were optimized within the training set by five-fold cross-validation using a Latin hypercube grid, with model selection based on cross-validated ROC AUC. The finalized models were then refit to the training data and evaluated on the held-out test sets, where accuracy, feature importance, and ROC AUC were computed.

To evaluate the extent to which models trained on one cohort generalize to the other, we implemented four cross-cohort validation training regimes in addition to standard random 75/25 train--test splits. These regimes were designed to systematically assess (i) cohort-specific internal validity, (ii) pooled internal validity, and (iii) cross-cohort generalizability. First, we trained and tested models within each cohort separately (BPRHS-only and PROSPECT-only). Second, we trained and tested models on the pooled integrated dataset (BPRHS + PROSPECT), allowing the algorithm to learn from the combined feature structure. Third, we trained models on the pooled integrated dataset but evaluated them within each cohort separately, by holding out a stratified test set from either BPRHS or PROSPECT while the remaining data (including the other cohort) formed the training set. Finally, we performed “pure” cross-cohort evaluation by training exclusively on one cohort and testing exclusively on the other (train-BPRHS/test-PROSPECT and train-PROSPECT/test-BPRHS). All cross-cohort splits preserved within-cohort class balance via stratification on diabetes status. These complementary training--testing regimes enabled us to describe how well models built on the harmonized feature set transferred between cohorts, and whether pooling the cohorts improved predictive performance in the smaller modeling dataset for PROSPECT.

#### Case weighting

RF models trained with case weighting provide a flexible approach for handling diverse, harmonized tabular datasets that combine multiple cohorts with different distributions, sizes, and outcome prevalences.

In our study, case weights were defined as the inverse of the class frequency within each cohort and supplied directly to the model during training. This design allowed the algorithm to balance the influence of participants from the larger BPRHS cohort against those from the smaller PROSPECT cohort, while also upweighting underrepresented diabetes cases relative to non-diabetes controls.

Unlike downsampling, case weighting does not discard data and therefore preserves the full range of heterogeneity in dietary, health, and social features. This is particularly important in integrated datasets where the distribution of predictors may vary between sites or populations, since retaining all observations ensures that rare but potentially informative patterns are not lost. By adjusting only the contribution of each row to the model fit, case weighting provides a principled way to equalize representation across imbalanced subgroups while maintaining model interpretability and comparability.

### SI Results: Random Forest model

#### ROC curves by training regime

The ordering of the ROC curves changed across the false-positive range (Figure 4C). At high specificity, the two regimes tested on PROSPECT (PROSPECT-only and Train Both → Test PROSPECT) lay below the BPRHS-only and pooled curves. As the threshold relaxed, the pooled-training model tested on PROSPECT improved more rapidly than the PROSPECT-only model and performed best of all regimes at moderate false-positive rates. At high false-positive rates, the PROSPECT-only curve rose above the BPRHS-only and pooled curves.

#### Feature importance

Beyond the leading features reported in the main text, the pooled and BPRHS-only models also ranked cholesterol, C-reactive protein, HDL and several FFQ features (alcohol, added and total sugars, fructose, sucrose and acesulfame potassium) highly (Figure 5A). In the PROSPECT-only model, the strongest predictors were plasma triglycerides, age, systolic blood pressure, waist circumference, LDL and BMI, followed by platelet count, C-reactive protein, creatinine and total cholesterol; the only SDOH feature ranked highly was the time a non-cohabiting child would take to reach the respondent in case of need.

#### Distributions of top-ranked predictors

The leading clinical and dietary predictors differed by diabetes status within each cohort, and levels were generally higher and more variable in BPRHS than in PROSPECT (SI Figure 1A–C). The time for a non-cohabiting child to reach the respondent differed little by cohort or diabetes status (SI Figure 1D). Among SDOH features (SI Figure 2), PROSPECT participants reported more social activities than BPRHS participants, 47% of BPRHS participants had less than a high school education, and most participants in both cohorts were born in Puerto Rico (BPRHS 96.6%, PROSPECT 87.4%).

#### Sensitivity to the imputation approach

To assess sensitivity to the imputation approach, we repeated the pooled V1 analysis on the same ten splits in two ways: with median imputation in place of k-nearest-neighbors imputation, and with missingness-indicator features added so that informative missingness could be learned. Neither changed test AUC or accuracy materially, the selected features overlapped closely with those of the main analysis, and the leading predictors were unchanged (SI Table 2). The results in the main text are therefore not sensitive to the choice of imputation method.

SI Table 2: Sensitivity of the Random Forest worked example to the imputation approach. Pooled BPRHS + PROSPECT model predicting diabetes status at V1; values are means over the same ten training–test splits (95% CI).

| **Imputation of numeric features** | **Test AUC** | **Test accuracy** | **AUC difference vs k-nearest neighbors** | **Feature overlap with k-nearest neighbors (Jaccard)** |
| --- | --- | --- | --- | --- |
| k-nearest neighbors | 0.778 (0.767,0.789) | 0.710  (0.701, 0.719) | — | — |
| Median | 0.778  (0.767, 0.789) | 0.715  (0.706, 0.723) | 0.000  (−0.003, 0.003) | 0.92 |
| k-nearest neighbors + missingness indicators | 0.776  (0.766, 0.786) | 0.711  (0.702, 0.720) | −0.002  (−0.004, 0.001) | 0.89 |

Only the imputation of numeric features differs between rows; categorical features were mode-imputed in all rows, and all other steps were as in the main analysis. Missingness indicators are one 0/1 feature per predictor marking a missing value, added before imputation. 95% CI: mean ± 1.96 × standard error across splits; AUC differences are paired within each split. Feature overlap: Jaccard index between the sets of Boruta-confirmed features, averaged over splits.

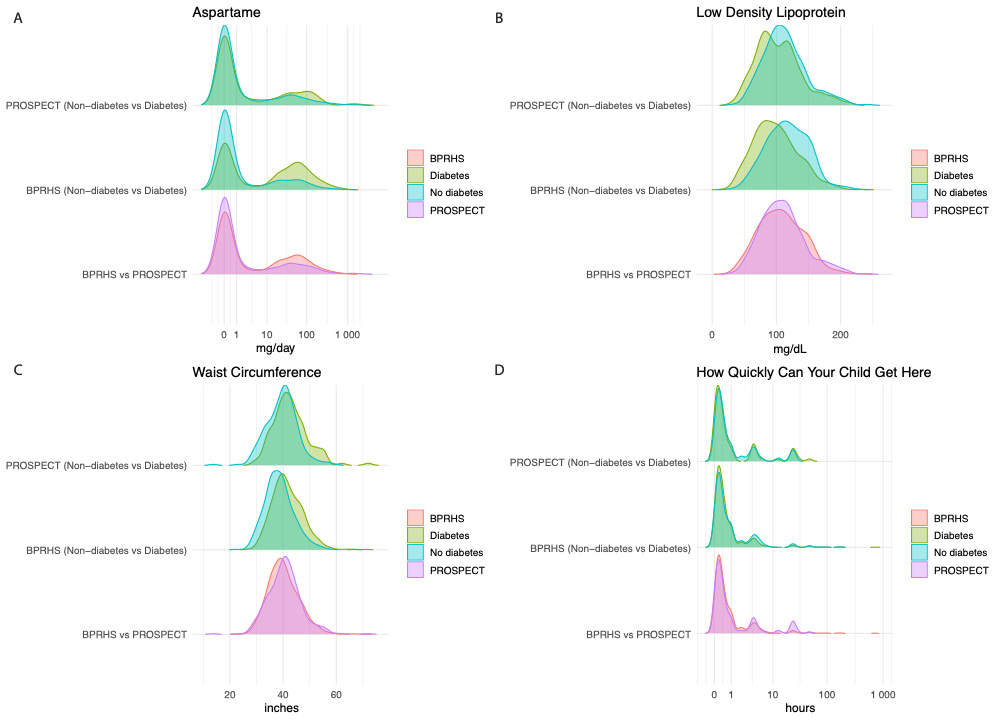

SI Figure 1: Distributions of selected predictive features from the BPRHS + PROSPECT Random Forest model stratified by cohort and diabetes status (0 = No diabetes, 1 = Diabetes). Panels show (A) aspartame intake, (B) low-density lipoprotein (LDL), (C) waist circumference, and (D) the time required for a non-cohabiting child to reach the respondent in case of need.

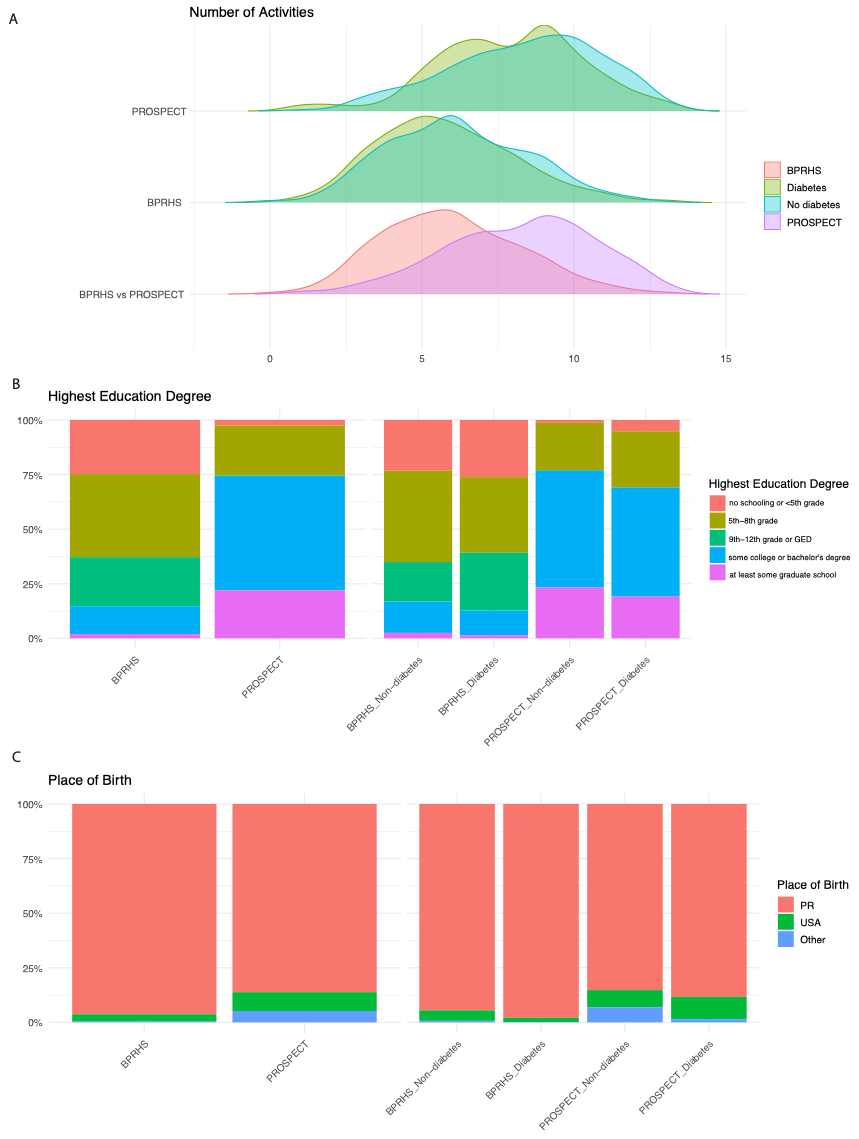

SI Figure 2: Distributions of the top-ranked social determinants of health (SDOH) features from the harmonized (BPRHS + PROSPECT) RF model, stratified by cohort (BPRHS, PROSPECT) and diabetes status (0 = No diabetes, 1 = Diabetes). (A) Number of activities in which the participant engages (B) Highest education level attained (C) Place of birth.
